# Process of Use of Evidence Products by Frontline Maternal, Newborn and Child Health Staff at the Facility Level in Ghana

**DOI:** 10.1101/2023.09.24.23296046

**Authors:** Gordon Abekah-Nkrumah, Doris Ottie-Boakye, Richmond Owusu, Johnson Ermel, Sombié Issiaka, Anita Asiwome Adzo Baku

## Abstract

Although the use of evidence products has been identified as beneficial in improving reproductive, maternal and child health, very little is known about the processes that facilitate the use of these evidence products by frontline Maternal, Newborn and Child Health and Reproductive and Child Health (RNCH/RCH) practitioners in decision-making on the issues that confront them in their line of work. This study explored the processes that facilitate the use of evidence products in reproductive, maternal and child health service provision in selected healthcare facilities in Ghana. Using a cross-sectional qualitative design, interviews of 24 frontline RNCH/RCH practitioners across 16 healthcare facilities in the Greater Accra, Eastern, and Ashanti regions of Ghana were conducted. The study employed a semi-structured interview guide modelled along the Action Cycle phase of the Knowledge-To-Action (KTA) framework, and the objectives of the study. Themes were built out of the transcribed data. The study revealed that the processes that facilitate the use of evidence products include problem identification activities (such as service evaluation, using accepted benchmarks, inadequate logistics for providing service and client feedback) and, adapting knowledge to their local context. The usual suspects of socio-cultural and health systems-related factors such as resource constraints and human factors were found to hinder the use of evidence products in providing RNCH/RCH services. The study contributes to knowledge by operationaliing the KTA, simplifying the concept of knowledge sustainability and makes it easy for adoption and implementation especially at the frontline. Appropriate interventions that rely on strong education to navigate the societal norms and beliefs that inhibit the uptake of evidence-based care by clients will be essential in improving the use of evidence to inform practice decisions.

## Introduction

The provision of quality maternal and child healthcare services is essential for reducing maternal, newborn and child morbidity and mortality globally. The World Health Organization (WHO) emphasizes the importance of evidence-based practice to improve maternal and child healthcare outcomes. Initiatives to improve maternal, newborn and child health (MNCH) and reproductive and child health (RCH) cut across several dimensions and include the Sustainable Development Goals (SDGs). The SDGs seek to reduce the number of maternal deaths per 100,000 live births globally to less than 70, reduce neonatal mortality to as low as 12 per 1,000 live births and under-5 mortality to at least 25 per 1,000 live births by 2030 (1).

Since the introduction of these ambitious goals, considerable progress has been made globally in terms of maternal and child health. In Ghana for example, the Ghana Maternal Health Survey 2017 estimated maternal mortality ratio as 310 per 100,000 live births, with under-5 and neonatal mortality reducing to 52 per 1000 live births and 25 per 1000 live births respectively (2). Likewise, approximately 89% of pregnant women had at least 4 antenatal clinic visits with skilled birth attendance reported as 79%. Despite the significant progress made within the past two decades, recent data suggests that maternal deaths contributed to 14% of all deaths in Ghana from direct and indirect causes (2,3). It is in the context of improving such adverse MNCH/RCH outcomes that the use of evidence products (existing research, policies, guidelines, protocols etc.) by frontline MNCH/RCH staff to inform decision-making is viewed as a step to improve quality and consequently cost-effectiveness.

Maternal healthcare staff play a critical role in ensuring safe and effective maternal care. The utilization of evidence products requires healthcare providers to incorporate the best available evidence with their clinical expertise and patients’ preferences. Nonetheless, research indicates that healthcare staff often rely on their personal experiences, thereby under-utilizing available evidence products when making clinical decisions (4–6). The use of evidence-based products provides benefits of standardized care, and scientifically proven evidence-based information to improve upon clinical decision-making and enhance the quality of care and outcomes for patients (7–9). Although evidence products have been proven to be beneficial and effective in guiding clinical practice, literature reports that frontline health workers underutilize them and, hence do not realize the associated benefits and impact (3–5), especially in terms of quality of care improvement (10–14). Previous studies have highlighted that selected sub-Saharan countries have limited use of clinical guidelines by healthcare workers [15-17].

Notwithstanding the low utlisation of evidence products for clinical decisions, existing research on evidence product utilization especially in the African context seems to focus more on policymakers at the national level rather than the frontline staff at the health facility level. For example, a search through the Ghanaian literature on MNCH/RCH evidence used by frontline health staff found only three papers in Ghana (15–17). Whereas the first paper (15) was on decision-making in general, the third paper (17) examined the determinants of evidence use by frontline MNC/RCH staff. Thus, except the second paper (16), there is currently no study that focuses on the process that facilitates the use of evidence products (existing research, policies, guidelines, protocols etc.) to inform decision-making by frontline MNC/RCH staff.

This paper therefore seeks to examine processes that facilitate the use of evidence products (existing research, policies, guidelines, protocols etc.) for care decision-making by frontline MNCH/RCH staff in selected health facilities. Specifically, the study:

- Examines the nature and importance of the use of evidence products by frontline MNCH/RCH staff in selected health facilities in Ghana.
- Examines processes that facilitate the use of evidence products by frontline MNCH/RCH staff in selected health facilities in Ghana to support practice decisions.

### Theoretical Framework

The current paper examines processes that facilitate the use of evidence products by frontline MNCH/RCH staff to support practice decisions. There are several models used to explain the process of translating knowledge (evidence) to support decision-making. The Knowledge-to-Action (KTA) Framework (see Fig 1) developed by Graham and colleagues (18) remains one of the most highly cited in the literature. The KTA framework is based on a system’s theory assumption, and it argues that knowledge producers and users operate in a knowledge system that is responsive, and adaptive but unpredictable. The KTA framework is divided into two distinct but related phases, namely; the Knowledge Creation phase and the Action Cycle phase (19). The Knowledge Creation phase acts like a funnel and puts knowledge into usable units, starting with knowledge inquiry (health research literature) to knowledge synthesis (systematic reviews, and meta-analyses) and knowledge products/tools such as best practice guidelines and protocols, clinical pathways or decision aids and videos (19). The Action Cycle of the KTA framework is iterative and involves steps needed for a unit of knowledge to reach widespread use to change behaviour and or attitudes. This includes the identification of a problem (i.e. identification, review and selection of knowledge), adapting knowledge to the local context (individual practice setting or particular patient), assessment of facilitators or barriers to knowledge use in a particular situation, selection and implementation of interventions to fit the local context, monitoring the use of knowledge, evaluation of outcomes and finally, implementation of systems to sustain the use of knowledge (18,19).

**Fig 1:**
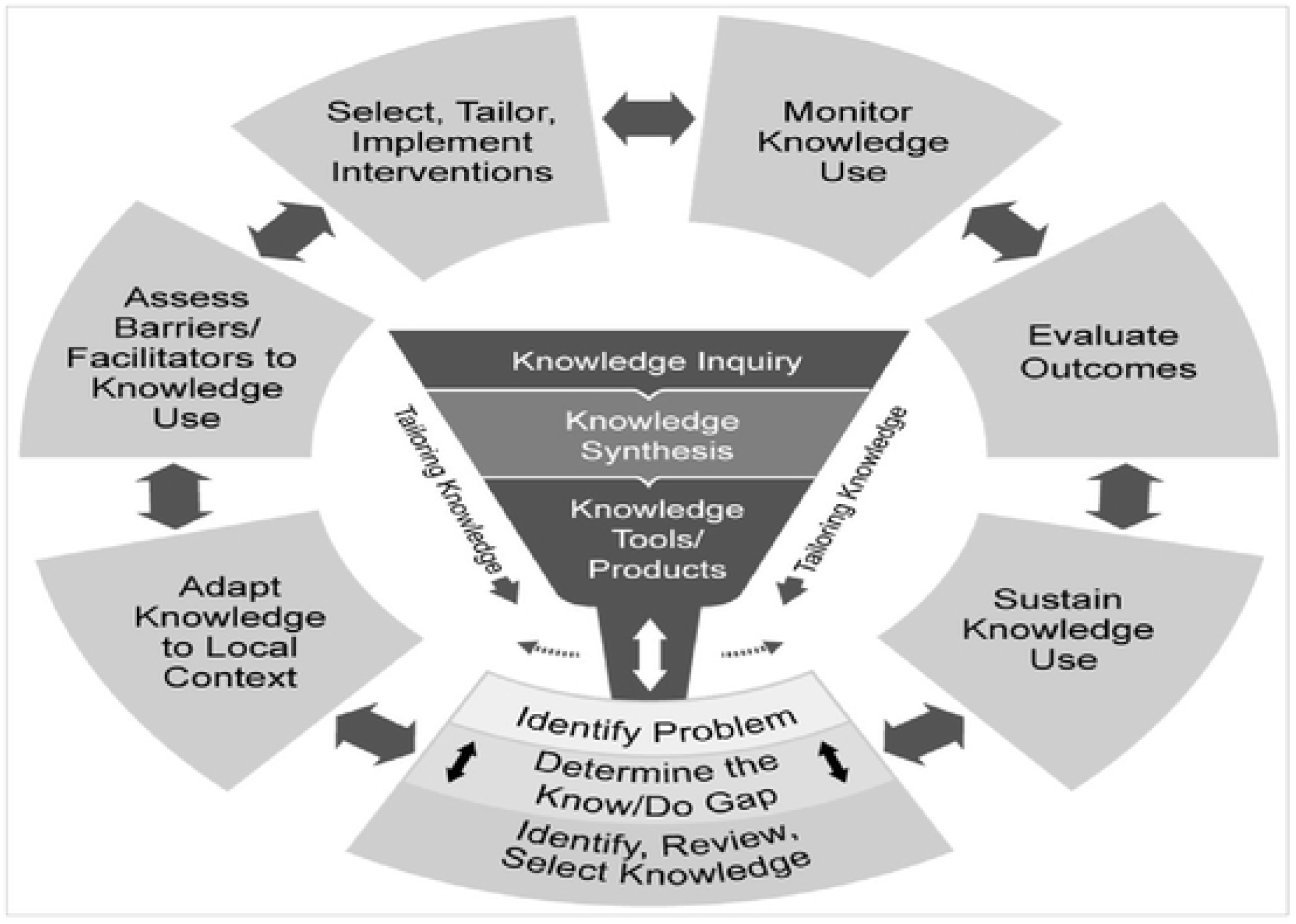
Knowledge to Action Framework. Source: adopted from Graham et al. (18)

Given that the study focuses on the use of existing evidence products to inform decision-making by frontline MNCH/RCH staff, we assume that knowledge already exists and that what constitutes a challenge is processes that facilitate the use of existing knowledge (i.e., research evidence, protocols, guidelines etc) to inform practice decisions. Thus, the use of the KTA framework in the context of this study is limited to the Action Cycle phase, starting from problem identification through to sustenance of knowledge use. The components of the Action Cycle phase are further discussed below.

#### Problem Identification

Although the Action Cycle is iterative, problem identification is normally recommended as the starting point for the deliberate application of knowledge to change behaviour and attitude. It compares current practice with existing knowledge to ascertain whether there is a practice gap that needs to be addressed(7,20).

#### Adapting Knowledge to Local Context

This step of the Action Cycle focuses on how existing knowledge could be organised to ensure that it is useful and appropriate to a specific circumstance (21,22). For example, it is argued that adapting existing clinical practice guidelines to fit local circumstances is not merely good for improving the relevance of the guidelines but also serves to improve ownership by end-users (23). Notwithstanding, there is equally the risk that adapted guidelines may deviate from the original evidence that informed the development of the guidelines (24).

#### Assessment of Barriers and Facilitators to Knowledge Use

This focuses on factors that influence (promote or constrain) the uptake of knowledge. Three main factors have been identified as influencing the use of evidence in the context of healthcare. For example, while Majid et al. (25) categorised the factors under knowledge and attitudes, Humphries et al. (26) classified the factors as information, organisational structure and processes, organisation culture and individual interactions. Other authors have classified such constraints or facilitators as related to organisational support and organisational change (27), personal attitudes, availability of resources and knowledge and skills (28) and attitudes and knowledge (29).

#### Select, Tailor and Implement Interventions

This is a three-part process that deals with the actual planning and implementation of an intervention (use of evidence) to bring about an intended change (30). The first part focuses on selecting effective interventions. However, given the possibility that different contexts may respond differently to the same interventions, tweaking interventions to fit the specific circumstances of different contexts can be key to success. The final part of this process is the actual implementation of the selected intervention. To ensure that the selected intervention produces the intended change, it should not only be effective but must also fit the specific circumstance of the context in question.

#### Monitoring of Knowledge Use

This component helps stakeholders to know whether the implementation of the interventions as discussed in the previous stage has resulted in the anticipated change. The monitoring focuses on three areas; whether there has been a change? The extent of the change and the factors accounting for the change. The monitoring can be done using robust scientific evaluation methods either qualitatively or quantitatively.

#### Sustain Knowledge Use

The importance of this phase is based on the assumption that successful interventions may not up-scale themselves unless systematic steps are undertaken to let it happen. This phase therefore focuses on taking systematic steps to ensure that positive changes occurring as a result of the implementation of the knowledge intervention are scaled up. It is suggested that factors such as perceived benefits and risks, relevance, leadership, policy integration, resources and politics are some of the factors that can aid the scaling up and sustenance of knowledge use (7,18,20,21).

### Methods Study Design

The study used a cross-sectional qualitative design and targeted frontline healthcare workers involved in the provision of MNCH/RCH services (medical doctors, nurses, midwives, public health officers, enrolled nurses, registered general nurses, and community health nurses) in 16 health facilities in 3 regions (Greater Accra, Ashanti and Eastern Regions). The health facilities were carefully selected to include those in rural and urban areas as well as areas that do not have demographic surveillance sites to reduce the possibility of contamination from the effect of demographic surveillance activities.

### Sampling Method

The choice of the Greater Accra, Ashanti and Eastern regions is based on the fact that they have the highest population of health facilities in addition to having different types (ownership and hierarchy) of health facilities. Using willingness to participate in the study, 16 health facilities were selected from the 3 regions. This includes (a) Ashanti: 1 Community Health Planning and Services (CHPS), 1 Health Center, 2 District Hospitals and 1 Regional Hospital; (b) Greater Accra: 1 CHPS, 3 Health Centers, 3 Regional Hospitals, 1 Tertiary Hospital and (c) Eastern: 2 Quasi-Government Hospitals and 1 District Hospital. From the 16 health facilities, 24 respondents were selected for the interview based on their willingness to participate and knowledge of the MNCH/RCH system in the respective health facilities.

### Data Collection and Analysis

A semi-structured interview guide was developed and used to collect data for examining processes that facilitate the use of evidence products by frontline MNCH/RCH staff to inform practice decisions. The interview guide contained specific questions that captured the socio- demographic characteristics of respondents and how key domains of the Action Cycle phase of the KTA framework (identification of a problem, adapting knowledge to the local context, assessment of barriers or facilitators to knowledge use in a particular situation, selecting and implementing interventions to fit local context, monitoring the use of knowledge, evaluating outcomes, and finally, implementing systems to sustain the use of knowledge) are deployed to facilitate the use of evidence products by frontline MNCH/RCH staff (18,19). The semi-structured interview guide is attached as supplementary material (S1 File).

All the interviews were carried out with the assistance of a trained interviewer and a field assistant. All interviews were audio-recorded, in addition to copious hand-written notes taken by the interviewer and her assistant. The audio recordings were transcribed word-for-word into a Microsoft Office Word document. The transcripts were compared with the hand-written notes and identified inconsistencies were corrected. The final transcripts were thoroughly reviewed by the interviewer and her assistant to ensure that the content of the transcripts was a true reflection of the answers or discussions with interview respondents. Using similarities and differences in respondents’ answers, information in the transcripts was grouped into broad themes and sub-themes in line with the objectives of the study. The broad themes and sub- themes formed the basis of what is presented in the findings.

### Ethical Consideration

Ethical clearance for the study was sought from the Ghana Health Service Ethics Review with clearance number GHS-ERC010/05/18. Additional administrative approval was taken from relevant regional and district Directors of Health Services as well as heads of participating health facilities. Informed consent was obtained from all the study participants. All participants provided written consent.

## Results

### Characteristics of Participants

Study participants were made up of 6 males and 18 females. The relatively higher number of females is because more females work in MNCH/RCH compared to males. In terms of profession, 2 public health nurses, 3 enrolled nurses, 2 community health nurses, 8 midwives, 5 senior/principal/nursing officers, 2 nutrition officers and 2 specialist obstetricians and gynaecologists participated in the study. Respondents also cut across various departments/units in the MNCH/RCH service delivery chain. These include 6 maternity/ANC wards, 5 RCH/family planning units, 3 cervical cancer screening units, 2 public health units, and 1 mother and baby unit. The rest include 2 respondents from the nutrition unit, 1 from the nursing unit, 2 from CHPS compounds and 2 consultants.

The findings are structured to respond to two main questions. First, what is the nature and importance of the use of evidence products by frontline MNCH/RCH staff, and secondly, processes that facilitate the use of evidence products by frontline MNCH/RCH staff to inform practice decisions.

### Nature and Importance of Use of Evidence Products

The section discusses the types of MNCH/RCH services offered in the health facilities interviewed, sources of MNCH/RC-related evidence products and the importance of using evidence products in practice decision-making as follows.

### Scope of MNCH services offered

Participants across the different levels of care irrespective of location (rural and urban) cited the key MNCH/RCH services offered by their health facilities as family planning services, postnatal care, nutrition care support, guidelines and rehabilitation, school and community health education, antenatal care services and delivery and labour services. Other services include cervical cancer screening, breast cancer screening, child welfare clinic services (CWC), adolescent health care services, sexually transmitted infection (STI) treatment and home visits. Neonatal and sick baby assessment and care, abortion care, and counseling were the least provided services. The findings indicate that the level of a health facility (primary care, secondary or tertiary) influences the type of MNCH/RCH service provided.

### Source of MNCH/RCH-related evidence products

Study participants rely on diverse sources for evidence products to inform MNCH/RCH practice. Nine participants mentioned training received either through in-service or on the job. About 10 of the participants mentioned the Ghana Health Service (GHS) as their main source of evidence products (guidelines, frameworks, algorithms and protocols) for use in the provision of MNCH/RCH services. Respondents also indicated that workshops, school, health facility-initiated meetings (weekly and monthly meetings) and the District Health Directorate (DHD) as their main sources of evidence products. Others indicated that their colleague workers and the use of models (dummies) constitute their main source of MNCH/RCH evidence products for their day-to-day decisions. Others mentioned development partners (UNICEF, WHO, JHPIEGO etc.). Ironically, only one rural participant cited online portals as one of the sources of MNCH/RCH-related evidence.

### Importance of evidence-based practice in MNCH/RCH service provision

Respondents described the use of evidence products in MNCH/RCH practice as important, giving varied reasons. They argue that evidence-based MNCH/RCH practice does not only benefit service providers but also users of services. They suggest that it serves as a source of health education and helps to streamline healthcare for clients. They acknowledged that it makes their work easier and also puts them in a position to do the right things in service delivery. To some, it serves as a guide for training service providers. They mentioned that evidence-based MNCH/RCH practice helps to monitor service delivery, educate clients on service provision and finally serves as a tool for identifying and managing risk. This is captured by a respondent below:

> *“They are very important because they serve as a guide to service providers, they also serve as a guide to monitor service delivery by the health service provider and also they serve as a guide for the training needs of health workers”* - (Gynecologist, Regional hospital, Urban area).

### Processes that Facilitate the Use of Evidence Products in Clinical Practice

In this section, we discuss processes that facilitate the use of evidence products by frontline MNCH/RCH staff based on the components of the KTA framework.

### Problem identification

As per the KTA framework, problem identification is about the identification of practice gaps to be addressed. The findings suggest that frontline MNCH/RCH staff work to identify practice gaps so as to improve their practice based on existing or new evidence. Problem identification is examined from the perspective of providers.

Respondents indicated that evaluation of the service provided, using accepted benchmarks, inadequate logistics for providing the service in question and clients’ feedback on service rendered helps to identify service and practice gaps. Respondents also acknowledged that other sources of information for identifying practice gaps include, facility-level service performance data via the strategic planning framework (i.e., setting of targets, monitoring and evaluation and examination of case outcomes). On inadequate logistics, a Community Health Nurse in charge of a Reproductive and Child Health unit averred:

> *“You are supposed to perform a procedure that requires that you have some logistics in place to be successful and yet not available. In that case, you should know that the outcome you seek you may not get it. This for me is a gap you need to address”* - (Community Health Nurse, Health Centre, rural area).

A nutrition officer in an urban hospital elaborated on the issue of monitoring and supervision in identifying gaps as follows:

> *“We go to the various Units to see if they are practicing what they have been taught. For instance, how to initiate breast feeding. We ask the mothers when we go to the ANC or the RCH unit. …if the mother is not able to breastfeed, we do ask, were you taught when you were pregnant or when you gave birth, were you helped to position the baby so that we will know whether what you taught the staff is being practiced or maybe it didn’t go down well.”*- (Nutrition Officer, Hospital, urban area).

Another study participant on the use of existing reports from documentation also highlighted: *“Whatever you do for a client, you document. So, per what the health personnel has documented, you will know that he/she is on course with the protocol or is deviating from the protocol”* – (Midwife, CHPS compound, rural area).

On the problem identification using client’s feedback, study participants reported that the condition of clients at visit, clients’ vital statistics, clients’ adherence to recommended service use, service outcomes etc. are key in helping them to identify knowledge or evidence gaps in their practice. The pathways to identifying gaps in the use of evidence in clinical practice were summarized by a respondent as follows:

On clients’ vital statistics, an enrolled nurse in an urban health center said:

> *“We do the tracing in the antenatal card when they visit every day. So, let’s say, the person came last month and her BP was 100/70, which of course is in a normal range, and then the weight was 90. Then the next month she comes, the BP has risen to say, 150/90 and then the weight may be has deteriorated to say 80 or 75, then we know there is a problem.”*- (Enrolled Nurse, Health Center, Urban area).

### Adapting knowledge to local context

Study participants averred that using knowledge or evidence products in their original form should be the gold standard. Nonetheless, there are context-specific challenges that sometimes require that existing or new evidence products (protocols, guidelines research evidence etc.) are tweaked a little to suit local context and therefore enhance ownership and usage, albeit that adaptation has its challenges. The statement below from one of the respondents confirms the fact that frontline MNCH/RCH staff engage in actions that seek to adapt existing and new knowledge to suit their context.

> *“…..What I’m trying to say is that it is not every case that the protocol works. Sometimes you have to adapt a standard on nutrition support care, other than that, the clinical outcomes for those cases will always be bad.”* - (Nutrition Officer, Hospital, Urban area).

It is important to emphasize that the extent to which one can adapt existing or new knowledge to the local context depends on the level of the health facility in question, and more importantly availability of resources. For example, a higher-level health facility with adequate staff, well trained on the protocol or guideline involved, will be able to adapt the protocol or guideline to their context better than a lower-level health facility without the required resources. This is captured by a respondent as follows:

> ***“****So, when I go for the training or whoever goes for it, when you come, you train the people that you work with and then you equip them with the knowledge and make it simple to implement in our facility. But sometimes you don’t have the people to do the training well for everybody and that can be a problem”* - (Senior Nurse Officer, Hospital, urban area)

Additionally, the level of teamwork and collaboration among the provider team as well as community engagements, to a large extent determine the level of knowledge adaptation. The two statements below capture how knowledge adaptation is accomplished through teamwork, collaboration and community engagement:

> *So, ideas are invited from all team members to draw the plan. After that, we just assign tasks to team members and later bring all our reports together. The reports help us to know whether we are being successful using the protocol or if there is a need to change something along the line - (Midwife, Hospital, Urban area)*
>
> *“We work with community health volunteers on home visits and outreach or immunization or weighing or education. To get them to do what is in the protocols, we educate them in a way that will make them understand so they can do the work we want them to do. Sometimes it means using things available in the community to help them to understand.”* - (Principal Community Health Nurse, Health Centre, Urban area)

### Barriers to evidence use

The findings suggest that there are inherent barriers to the use of evidence products by frontline MNCH/RCH staff in making daily practice decisions. These include client-related and health systems-related barriers which have been discussed in detail below.

### Client-related barriers

Client-related factors that constrain the ability of frontline MNCH/RCH staff to use evidence products in their practice decisions include socio-cultural (societal norms and belief systems) and economic factors (access to resources and education). According to respondents, societal beliefs and norms manifest in ways such as women needing permission from their husbands or in some instances fear of accepting to utilize services crucial to securing better health outcomes for them. The quote below suggests that societal norms (e.g., a husband’s authority) are crucial to administering evidence-based life-saving interventions by frontline MNCH/RCH staff.

> *“Most people when they come, you can see that it’s an emergency and you have to quickly attend to them. Some women will tell you that unless my husband comes, I can’t open myself naked before you. This is normal because there are some religious organizations and cultural beliefs that if you want to render the service if you want to even touch a client’s private part, the husband will have to be there to give consent. So, in that case, maybe if a protocol is given, do this and do that, you will realize that you will have to miss the first or second step”*- (Public Health Nurse, Hospital - urban area).

Poverty and low levels of education among clients also play a key role in the ability of frontline MNCH/RCH staff to utilize evidence in daily clinical decisions. The respondents explained that poverty creates affordability challenges (especially for medicines and diagnostics) making some clients opt for alternatives other than what has been prescribed for them by the MNCH/RCHS staff as per the quotes below.

> *“The challenge we have is sometimes the women are not able to afford the laboratory services like the lab test, the screening and the Pap Smear. They don’t have the money to do it. Most of them come and say “I don’t have money, I will go and come”* – (Midwife, Hospital, Urban area).

Additionally, the respondents suggest that lower levels of education in the context of strong cultural beliefs manifest in fear and resistance to the adoption of evidence-based advice. For example, clients with non-communicable diseases like hypertension after receiving health education may not adhere to their medication regime, while in other cases, women may opt for family planning methods that may be detrimental to their health per available evidence. The quote from a public health nurse below explains this fact.

> *“Sometimes the person is pregnant. You educate the person to eat those food items that you the health worker think are nutritious and will help the baby. But that is the same thing that the family, per their belief, thinks that a pregnant woman consuming such food items will give birth to an abnormal baby. So, these women are scared to consume such food items”* - (Public Health Nurse, Hospital - Urban area)

### Health systems-related barriers

Several health system factors as discussed below were suggested by respondents as constituting barriers to the use of evidence products by frontline MNCH/RCH staff.

#### Poor staff attitude towards the use of protocols

One of the health systems barriers indicated by study participants was poor staff attitude towards the use of protocols in providing MNCH/RCH services. Although health personnel have received training on the use of protocols for routine clinical care, some do not follow the procedure in service provision. In a hospital located in an urban area, one Senior Nursing Officer said:

> *“Although you’ve taught everybody to go through the protocol, people will still not use it and will want to do whatever they want. They want to finish whatever they want to do quickly, either they are in a hurry, or they think it’s not necessary to go through the protocol*” - (Senior Nursing Officer, Hospital, Urban area).

#### Inappropriate use of protocol

This refers to cases where protocols are available in the facility but may not be accessible to those who may need them for their work. A Senior specialist Obstetrician and Gynecologist in an urban hospital pointed out:

> *“Sometimes, some of the protocols are available and may be sitting in somebody’s office who is not the direct user. A good example is the standard treatment guideline, supposed to be in the consulting rooms. Some prescribers don’t have it and are neither trained on its use nor yet are supposed to use it. So that can be a barrier to its proper use.”* - (Senior specialist Obstetrician and Gynecologist, Regional hospital, Urban area).

#### Resource constraints

The use of evidence products to inform practice decisions requires resources. There are instances where protocols are made available without accompanying resources to make their use efficient and effective. This is not limited to urban facilities but also in rural settings. A Senior Specialist Obstetrician/Gynecologist emphasized:

> *“Some of those protocols may require change of working tools, equipment etc. I believe that, well, we are trying but it can be better, it’s not completely there sometimes”* - (Senior Specialist Obstetrician and Gynecologist, Regional Hospital, Urban area).

#### Limited staff strength

Inadequate health personnel to provide MNCH/RCH services was also cited as a barrier to the use of evidence by frontline MNCH/RCH staff in both rural and urban facilities. According to them following protocols, guidelines and new evidence sometimes require not only time but also the requisite personnel. They argue that the challenge is even aggravated when they often work below the required staff norms in a time of increasing utilization. In this direction, a Public Health Nurse cited:

> *“We are a big health facility with few staff. If you want to follow the protocols strictly, it takes time and the clients grumble. So, staffing is a problem”* - (Public Health Nurse, Hospital - urban area).

#### Conflict of MNCH/RCH protocols with other policies

Given the numerous protocols in MNCH/RCH care, the study found that some protocols are in sharp contrast with other existing healthcare policies. This puts service providers in a dilemma in the use of such evidence products to provide MNCH/RCH services to clients. One Midwife in a rural CHPS compound recounted:

> *“When somebody comes to deliver, our protocol says you should give antibiotics to prevent any infection due to the issue of the placenta. If such a client comes with insurance (NHIS card), the insurance policy says that the level of the facility, (i.e., CHPS zone) does not administer antibiotics. But, here in our CHPS zone too, people come here to deliver. So, what do we do?”* – (Midwife, CHPS compound - Rural area).

#### Inadequate training

Inadequate training was also cited as a barrier to the use of evidence in MNCH/RCH service provision. This they argued makes it difficult for health personnel to grasp the concepts in the protocols, guidelines etc. before their introduction. One study participant reported:

> *“What I see as some of the barriers is inadequate training for health personnel to be acquainted with protocols, guidelines etc. early enough before their introduction. Not that they don’t do the training, but the training should be as early as possible before the introduction so that people can start using the protocol.”* – (Senior Specialist, Obstetrician/Gynecologist, Hospital - urban area).

#### Overconfidence

Some study participants believe that some MNCH/RCH service providers are over-confident given their familiarity with numerous protocols. This could also be a barrier. One Midwife in an urban hospital said:

> *“It may happen at different places, due to familiarity, they think they already know”* - (Midwife, Hospital, Urban area).

#### Staff rotation

Another health system-related barrier mentioned by participants is the periodic rotation of health personnel from one unit to the other within the health facility. This becomes a challenge, given that one needs time to go through the protocol and grasp it. A senior Nursing Officer acknowledged:

> *“One of the challenges is periodic reallocation of health professionals from one unit to the other. So, initially, when they come, it becomes a challenge for the child unit. You need to take them through the protocol, and it takes time before they grasp it.”* - (Senior Nursing Officer, Hospital, urban area).

Notwithstanding the barriers enumerated above, respondents were almost unanimous that they work hard to use evidence in their daily clinical decisions and that the quest for positive service outcomes, staff motivation or incentives, and in some instances the need to avoid unnecessary litigation constitute powerful incentives that keeps them persevering in the face of limitless constraints. This is well articulated in the quote from a respondent below.

> *“What motivates us is that if there is any problem and you know that you did what you’re expected to do and therefore the law would let go of you. But if you don’t do it and there is a problem, then you can be held accountable for it.”* - (Senior Nursing Officer, Hospital, Urban area).

### Selecting, tailoring and implementing interventions

The selection of evidence products (i.e., protocols guidelines etc.) is determined by the national level and cascaded through the different levels of the health system, with relevant training and logistics needed for implementation at the different levels. It is important to emphasize that detailed plans for implementing such evidence products are prepared at the health facility level in consultation with the community to ensure that implementation of the evidence product in question will be smooth and acceptable to encourage uptake. Using family planning, for example, participants narrated how new evidence products were selected, tailored and implemented. They indicated that a particular evidence product may for instance come from a Development Partner such as WHO or UNFPA and will first be adopted by GHS. During the adoption process, people with experience are tasked to look at the protocol and adapt it to suit local use, taking into consideration the kind of health services provided at each level. At each level of service provision, the protocol states what is expected to be done and the resources required. The adapted evidence product is then reproduced and disseminated in addition to training for those who will be using the evidence product in service provision. The trainer of trainer’s approach is often used such that a few super users are trained at the regional level, who in turn train others in the districts, and cascade it to the health facility level.

After the training, the next level is the implementation of the evidence product. Participants interviewed suggest that the process is normally preceded by a meeting of the personnel supposed to be working with the evidence product. The meeting is normally to gather ideas that will make it relatively easy to implement the evidence product in a manner that works in the health facility. Following the meeting, detailed plans are put in place including assigning duties to health workers in the health facility. This is done in line with the objectives of the facility as well as plans put in place to monitor outcomes of the implementation, such that identified gaps can be identified and addressed.

### Monitoring Use of Evidence

The study found that varied mechanisms are employed in monitoring the use of evidence products in MNCH/RCH service provision. These include the use of positive health outcomes, observations, and meetings (weekly, quarterly and monthly), the use of assessment indicators, either from clients’ or health personnel’s perspectives and the use of checklists for supervision. Other strategies recounted by participants were on-the-job training, reports (daily/monthly ward state), supervision (physical or supportive), mortality audit and research. Selected quotes from respondents have been reproduced below to emphasize this assertion.

> *“The diseases we are immunizing against we are doing surveillance on it. So, if less cases being reported of those diseases then it means the immunization is working and by extension, the protocol being used.”* - (Midwife, CHPS, Rural area).
>
> *“We have a checklist, and the supervisors do both physical monitoring and supportive supervision. So, when a protocol is introduced, you go from time to time to check whether staff are following it. This is done through some form of on-the-job training to ensure that they use it appropriately.”* – (Senior Specialist, Obstetrician/Gynecologist, Hospital - Urban area).
>
> *“We have our daily wards state every morning that helps us to know whether we are making progress or not. For instance, if there is a mortality incidence, we will carry out an audit to find out what really went wrong. Was it our fault or was it the fault of the mother or something really went wrong? In such audits, we are thorough and also take note of where we went wrong … and then, next time we are cautious about it.”*- (Senior Nursing Officer, Hospital - Urban area).

### Sustaining Knowledge (Evidence Products) Use

The study revealed that several approaches were used to sustain evidence of products use in MNCH/RCH service provision. Three themes emerged and these were empowerment, consultation and staff cooperation.

#### Empowerment

Study participants identified empowerment (teaching, sharing of ideas, training and continuous practice) as one of the key components of sustaining the use of evidence products in the provision of MNCH/RCH services. On the issue of teaching, a Principal Community Health Nurse in a rural health center had this to say:

> *“As I said, we go to the workshop, we learn it and come back and then teach it.”*

(Principal Community Health Nurse, Health Centre, Rural area).

Recounting her experience on sharing of ideas, an Enrolled Nurse in an urban health center echoed:

> *“When they go for workshop, when they come, we all sit down, and we share their ideas. So, the next time you also get a patient, you will also follow the same procedure, since it’s in line with the protocol that we have here”* - (Enrolled Nurse, Health Centre, Urban area).

Shedding more light on continuous practice as a way of sustaining the use of evidence, a Principal Nursing Officer in an urban hospital averred:

> *“With time, practice makes perfect. If we are doing it gradually, we’ll get there. But if you are not using it at all, you wouldn’t know what it even entails, and you may forget some of the things in the protocol. So, I think continuous practice helps.”* – (Principal Nursing Officer, Hospital, Urban area).

#### Consultation

Another key factor that helps to sustain the use of evidence products in MNCH/RCH service provision cited by participants is consultation. The study established that consultation is done through weekly, monthly or quarterly meetings or workshops where issues are discussed, and knowledge is impacted through consensus building devoid of imposition. Sharing her experience in meetings, a Public Health Nurse in an urban hospital said:

> *“When there is an emergency issue to be discussed, we call them. We also have scheduled meetings, like quarterly or monthly meetings. Then we discuss the issues. We don’t impose anything on anyone. We all share our knowledge on how and ways to improve.”* – (Public Health Nurse, Hospital, Urban area).

#### Staff cooperation

The study identified cooperation of health personnel through teamwork, good relationships and firmness as a strategy to sustain evidence products use in MNCH/RCH service provision. Participants argued that good relationship and teamwork creates the environment for sharing acquired knowledge/ideas and experiences among frontline staff. This sustains the use of evidence products in providing services. Again, being firm by insisting that frontline staff do the right thing, ensures protocol adherence and strengthens knowledge use in MNCH/RCH service provision. Participants said:

> *“Sometimes we come together and talk about issues that constitute a common challenge. For example, someone goes to a workshop on preeclampsia, she will discuss what she learnt with those of us not at the workshop so that we will also learn some of what she learnt. If she also has anything new, she will discuss with us so that we will all know and do the work together.”* - (Midwife, Health Centre, Urban area).

The quote below from a nutrition officer from an urban hospital also emphasizes the issue of insisting that frontline staff make it their duty to do the right things first.

> *“In fact, I try to be very firm. So, you have to be firm. You will sometimes be insulted, but once the insult will not kill someone’s child, I’m ok. If you insult me and the right thing is done and the child is ok, my heart is happy”* - (Nutrition Officer, Hospital, Urban area).

## Discussion

The paper examined the nature of evidence products used by the frontline MNCH/RCH staff for practice decisions, their importance and the processes that facilitate their use. From the findings, evidence products in use cover a wide range of areas (family planning, postnatal, nutrition, antenatal, neonatal, adolescent health etc.) and were obtained from sources such as training, workshops, periodic facility-based meetings, schools, colleagues workers, online resources, development partners and parent organizations such as the Ghana Health Service and District Health Directorates. More importantly, frontline MNCH/RCH staff see the use of evidence products in practice decisions as essential not only for service providers but also for service users. Although there is no evidence to suggest that frontline MNCH/RCH staff follow the action cycle phase of the KTA framework in the same sequence as prescribed in theory, the findings nonetheless, suggest that it constitutes a relevant theoretical model for understanding processes that facilitate the use of evidence products by frontline MNCH/RCH staff to inform practice decisions.

From the findings, frontline MNCH/RCH staff have put in place processes (evaluation of services, resources and feedback) to identify gaps that need to be addressed as suggested in the literature(7,20). These findings also indicate that frontline MNCH/RCH staff in the health facilities studied have adopted mechanisms to adapt evidence products and make them relevant to their context as prescribed in the KTA framework and other studies (21–23). The findings however suggest that such adaptions may not be straightforward as argued in theory (24). Depending on the context, there may be resources as well as administrative and community-level political implications for such adaptations. Thus, depending on the capacity of the health facility to navigate these constraints, such adaptations could be extremely difficult. This to some extent may reflect the caution by Fervers et al. (24) that adaption of evidence products to local context can result in a deviation of practice from the original evidence.

Additionally, the existing literature suggests that barriers to the use of evidence products emanate mostly from organisational systems (25,28,29). For example, barriers have been operationalized to reflect knowledge and attitudes (25), organizational support and organizational change (27), personal attitudes, availability of resources and knowledge and skills (28,29). Consistent with these studies, the findings suggest that health system barriers such as poor staff attitudes, resource constraints, limited staff numbers and skills, conflicting policies and inadequate training, constrain the use of evidence products by frontline MNCH/RC staff in their practice decisions. More importantly, the findings suggest that besides health system factors, there are client-level factors such as societal norms and belief systems, poverty and education that strongly constrain the ability of frontline MNCH/RCH staff to deploy evidence-based practice. These findings can be crucial in informing the way interventions are deployed, knowing that strengthening health systems alone may not be enough to improve the use of evidence products by frontline health personnel.

Also, the findings of the study suggest that the selection and tailoring of evidence products are done mainly at higher levels of the administrative structure and that it is the implementation that is amenable to local-level participation and changes. This to some extent is different from the KTA framework, which argues that due to variation in context, it is appropriate not only to select but also to tailor interventions to suit local conditions to limit barriers to implementation (30). On this basis, one will expect that the evidence products selection process will reflect substantial input from those at the lower levels who have a better understanding of the context. This will ensure that both the selection and tailoring reflect local conditions to reduce to the barest minimum possible implementation barriers. There are several examples in the policy implementation literature where the need for substantial collaboration with all stakeholders especially those at the bottom shas been advocated as a strategy for improving the implementation of policies (31–34).

Theoretically, the essence of monitoring is to determine whether a change has taken place as a result of the use of evidence products, and if so, the extent of the change and the factors accounting for the change. Consistent with the KTA framework, the findings suggest that frontline MNCH/RCH staff deploy different mechanisms (health outcomes, observations, periodic meetings, use of assessment indicators either from clients or health personnel’s perspectives, and the use of checklists for supervision etc.) to monitor the use of evidence products in practice decisions. Most importantly, the findings suggest that the monitoring system as per the requirement of the KTA framework makes it possible for evidence product users to identify the extent of change occasioned by the use of evidence products. What is however not clear from the findings is whether the system deployed systematically collects data to help frontline MNCH/RCH staff to know the extent of change if any, and the factors responsible for same. For example, to what extent does the change fulfil the requirements of acceptability, appropriateness, fidelity, cost-effectiveness, penetration and sustainability as advocated for in the literature (35,36)?. Also, the findings do not suggest that in practice, evaluation is designed to specifically measure structural, process and outcome variation as articulated in the KTA framework. This constitutes a weakness in the evaluation mechanism deployed by the health facilities studied, given that any targeted intervention to address adverse outcomes may be difficult, if not impossible.

Finally, empowerment (ideas sharing, teaching, continuous practice), consultation (meetings, workshops, consensus building) and staff cooperation (teamwork, relationship building, firmness) approaches used by frontline MNCH/RCH staff to ensure sustainable use of evidence products in practice decisions is similar to the parameters of the original model by Graham et al., (18) as well as other applications of the original model (7,21,22). These models emphasised parameters (perceived benefits and risks, relevance, leadership, policy integration, resources and politics) that can help in scaling up knowledge use. Unlike empowerment, consultation and staff cooperation, the parameters emphasised by Graham et al., (18) are complex and may not be easily appreciated by practitioners and policymakers. Thus, it can be argued that the current study findings do not only offer a better operationalisation of existing frameworks for ensuring sustainable use of evidence products but also make them easy to appreciate and use, especially by practitioners and policymakers. For example, advocating for ideas sharing, teaching, continuous practice, consensus relationship building and firmness as a strategy to sustaining the use of evidence products among frontline health staff is straightforward to identify with, compared with terms such as relevance, leadership, policy integration, resources and politics.

## Conclusion

The study findings suggest that the use of evidence products is a pathway to improving maternal and child health outcomes. Thus, putting systems in place to strengthen the use of evidence products, especially at the health facility level is expected to not only consolidate the gains made over the last two decades but can also constitute a platform for seeking further improvements in outcomes.

The study findings also make clear the need to address challenges within the knowledge utilisation process and thus, provide clear adaptation guidance to prevent difficulties that arise from the complexity of the process. There is also the need to ensure that the process for selecting and tailoring knowledge is broad-based and benefits from substantial inputs from the lower levels to enhance relevance and adoption. Additionally, there will be the need to improve the monitoring framework to enhance acceptability, appropriateness, fidelity, penetration, cost-effectiveness and sustainability as advocated for by the KTA framework.

In addition, the study better operationalizes the KTA requirement for sustainability. For example, using concepts such as empowerment (ideas sharing, teaching, continuous practice), consultation (meetings, workshops, consensus building) and cooperation (teamwork, relationship building, firmness) simplifies the concept of knowledge sustainability and makes it easy for adoption and implementation, especially at the frontline. Besides the normal health systems factors that have been the focus of the knowledge translation and utilisation, individual (client) level factors are equally essential in addressing barriers to the utilisation of evidence products especially at the health facility level. Thus, interventions aimed at scaling up the use of evidence products in practice decisions by frontline staff should not only be aimed at addressing health systems challenges but also challenges at the individual level. In this direction, interventions that rely on strong education to navigate societal norms and beliefs that inhibit the uptake of evidence-based care by clients will be essential in improving the use of evidence products to inform practice decisions by frontline MNCH/RCH staff.

## Data Availability

Detailed data based on which the current paper was written is captured in the original report which was submitted to West African Health Organisation (WAHO). The raw data that was collected from the field and formed the basis of the original report is also the property of WAHO. Both the original report and the raw data can be made available upon request from WAHO.

## Acknowledgements

The authors acknowledge the immense support of WAHO in carrying out the study and the report from which this paper was developed. We are also grateful to all who participated in this study.

## Author Contributions

Conceptualization: GA, JE, SI, Data curation: GA, DO, RO Formal analysis: GA, DO, AAAB, Funding Acquisition: GA, Investigation: GA, DO, AAAB, SI, JE, Methodology: GA, DO, JE, SI, AAAB, RO. Project Administration: GA, RO Supervision: GA, Validation: GA, RO, Writing - original draft preparation: GA, RO, DO, Writing- review & editing: GA, RO, DO, JE, SI, AAAB

